# Automated epilepsy and seizure type phenotyping with pre-trained language models

**DOI:** 10.64898/2026.02.11.26346003

**Authors:** Ellie Chang, Kevin Xie, Daniel J. Zhou, Jacob Korzun, Erin C. Conrad, Dan Roth, Brian Litt, Colin A. Ellis

## Abstract

**Background:** Epilepsy is a common neurologic disorder characterized by recurrent, unprovoked seizures. Epilepsy manifests as different seizure types and epilepsy types, which have important implications for treatment and prognosis. Electronic health record systems containing longitudinal data on large epilepsy cohorts can be valuable resources for clinical research. However, detailed epilepsy phenotypes are poorly captured by structured data such as diagnostic codes and are instead buried in unstructured clinical notes.

**Methods:** We evaluated two transformer-based language models for automated epilepsy and seizure type phenotyping from clinical notes: a fine-tuned BERT model and a large language model, DeepSeek-R1. A subset of notes was annotated by epileptologists, and model performance was benchmarked against expert agreement. The best-performing model was then deployed across all epilepsy progress notes at a large academic medical center to generate patient-level longitudinal epilepsy and seizure phenotypes.

**Results:** Both models achieved performance comparable to expert agreement for classifying epilepsy type as focal, generalized, or unspecified (Matthews correlation coefficient [95% CI]: DeepSeek = 0.85 [0.80-0.90], BERT = 0.73 [0.67-0.80], human = 0.77 [0.70-0.83]) and classifying seizure type as convulsive or non-convulsive (DeepSeek = 0.74 [0.66-0.81], BERT = 0.60 [0.49-0.69], human = 0.49 [0.39-0.59]). For more granular classification tasks, DeepSeek maintained performance comparable to expert agreement, whereas BERT performance declined. Deploying DeepSeek-R1 on 77,049 clinical notes from 18,566 patients revealed system-level clinical patterns, including diagnostic stabilization over time, frequent co-occurrence of seizure types, and variation in seizure outcomes by epilepsy type.

**Conclusions:** By extracting expert-level epilepsy phenotypes from routine clinical text at scale, this approach transforms unstructured EHR data into a resource for longitudinal, population-informed epilepsy care. Automated phenotyping enables analyses of epilepsy trajectories and treatment outcomes that are not feasible with structured data alone, supporting future clinical and translational research applications.

## 1 Introduction

Epilepsy affects more than 60 million people worldwide and is characterized by recurrent, unprovoked seizures that vary widely in etiology, clinical presentations, and response to therapy. In routine clinical practice, decisions about treatment, counseling, and referral for surgical or device-based interventions depend critically on accurate classification of epilepsy type and seizure type. Misclassification or incomplete characterization can delay appropriate therapy, hinder surgical evaluations, and ultimately worsen patient outcomes. These distinctions also have direct implications for patient safety. Bilateral tonic-clonic seizures are the major risk factor for Sudden Unexpected Death in Epilepsy (SUDEP), which accounts for over 3,000 deaths annually in the United States, and SUDEP risk increases with the frequency of these seizures.^1^

Despite their central role in care, epilepsy syndromes and seizure types are poorly captured in structured electronic health record (EHR) fields like diagnostic codes. Instead, most clinically meaningful phenotypic detail is embedded in unstructured clinical documentation such as progress notes and EEG reports, where it remains inaccessible to large-scale analyses. As a result, although health systems now routinely collect longitudinal data on tens of thousands of people with epilepsy, most research cohorts still rely on small, manually curated datasets, limiting generalizability and constraining analyses of longitudinal outcomes such as seizure freedom, treatment response, and SUDEP risk.

Advances in natural language processing (NLP) offer a potential solution. Earlier rule-based clinical NLP systems require substantial domain expertise, are brittle to variation in terminology and documentation style, and scale poorly across heterogeneous corpora. In contrast, transformer-based language models, introduced in 2017,^2^ leverage self-attention to capture long-range contextual relationships and have demonstrated strong performance across clinical phenotyping, risk prediction, and temporal event extraction tasks, even with limited labeled data.^3^ These capabilities are particularly relevant in epilepsy, where phenotypic information is abundant but distributed across lengthy, narrative clinical notes.

Prior work has established the feasibility of applying NLP to epilepsy-related tasks. For example, Xie et al. (2022) fine-tuned BERT-based models to extract seizure outcome variables – including seizure freedom status, seizure frequency, and date of last seizure – from clinical notes, achieving performance comparable to expert annotators.^4^ Subsequent studies extended these methods to large language models (LLMs), reporting strong performance on selected epilepsy information extraction tasks, including seizure frequency, seizure type, and anti-seizure medication identification.^5–7^ However, prior work largely focused on a limited set of outcomes rather than systemically mapping fine-grained epilepsy syndromes and seizure types at health-system scale, and few studies have directly benchmarked LLMs against epileptologists for these nuanced phenotypes.

Building on prior work, we developed and evaluated automated methods for fine-grained epilepsy and seizure type phenotyping from unstructured clinical notes. Our aims were to validate model performance against board-certified epileptologists and to deploy the best-performing model to generate a longitudinal, health-system-wide corpus of epilepsy phenotypes. We evaluated a fine-tuned BERT^8^ model and an autoregressive LLM (DeepSeek-R1-Distill-Llama-8B^9^) under zero-shot and few-shot configurations, benchmarked them against expert annotations, and then deployed the selected model across nearly 80,000 clinical notes from over 18,500 patients who had seen an epileptologist between 2011 and 2024.

## 2 Materials and Methods

### 2.1 Data Collection

The source dataset was the text of outpatient notes from the epilepsy clinics at the University of Pennsylvania from 2011 to 2024. We previously extracted seizure freedom and seizure frequency from these notes using a validated transformer-based language model.^10^

### 2.2 Data Annotation

To develop and evaluate transformer models for epilepsy type and seizure type phenotyping, we first constructed an annotated dataset of notes. For each free-text note, we extracted the epilepsy and seizure phenotypes described within it. Specifically, for each note we asked:

A. “What type of epilepsy does the patient have?”
B. “What types of seizures has the patient had?”

Annotators assigned the most granular epilepsy and seizure types present in each note (the “all-way” scheme). For model evaluation, these specific labels were also mapped into broader category schemes as summarized in **Figure 1**. Epilepsy type (question A) was treated as a single-label classification task under all three schemes, with each note assigned exactly one label. For seizure type (question B), the binary and three-way schemes were treated as single-label tasks, whereas the all-way scheme was treated as multi-label classification, allowing each note to be assigned multiple seizure types.

**Figure 1.**
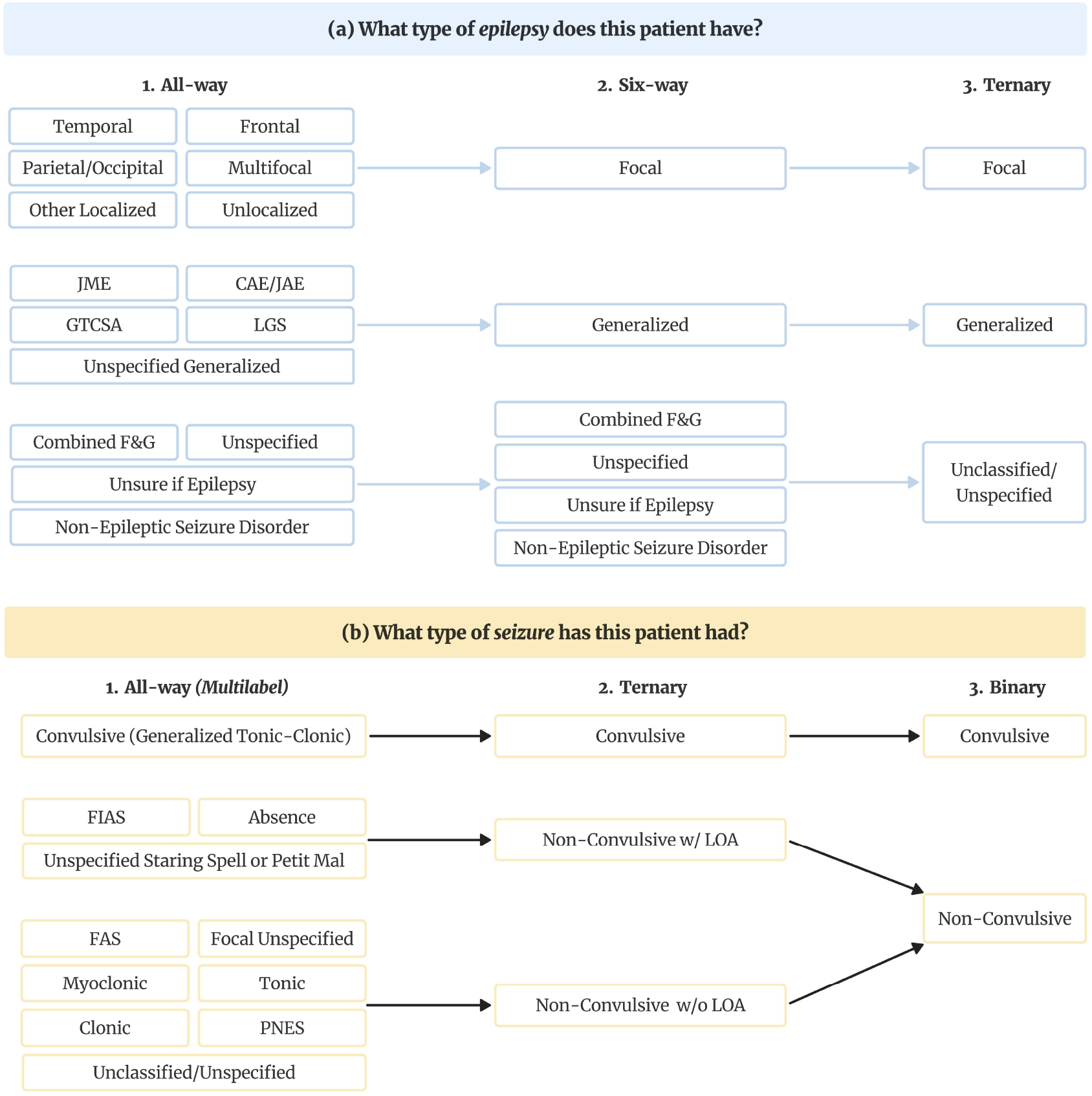
Epilepsy and seizure type classification schemes. (a) Epilepsy type labels were annotated at the most granular (“all-way”) level and subsequently mapped to six-way and ternary schemes for analysis. (b) Seizure type labels were similarly mapped from granular all-way categories to ternary and binary schemes. For binary and ternary seizure-type schemes, each note was treated as single-label with the most severe seizure type present in the note selected as the label. Only the all-way seizure-type scheme was evaluated as a multi-label task, allowing multiple seizure types per note. ***Abbreviations:*** *JME, Juvenile Myoclonic Epilepsy; CAE/JAE, Childhood/Juvenile Absence Epilepsy; GTCSA, generalized tonic-clonic seizures alone; LGS, Lennox-Gastaut Syndrome; F&G, Focal & Generalized; BTC, bilateral tonic-clonic; GTC, generalized tonic-clonic; FIAS, focal impaired awareness seizure; FAS, focal aware seizure; PNES, Psychogenic Non-Epileptic Seizures; LOA, Loss of Awareness*.

Three board-certified neurologists annotated 309 free-text clinic progress notes from the source dataset using the INCEpTION platform.^11^ Each note was independently labeled by two annotators, with disagreements adjudicated by a three-author consensus panel (EC, KX, CAE). Final labels for epilepsy and seizure types were merged into separate “ground truth” datasets. These fine-grained labels were then mapped to broader categories for higher-level classification tasks.

### 2.3 Model Development and Implementation

We fine-tuned and evaluated two transformer-based NLP models: a fine-tuned masked language model, BERT, and an autoregressive large language model, DeepSeek-R1 (**Figure 2**). BERT was trained using domain-adaptive pretraining on ∼64,000 unannotated clinical notes, followed by supervised fine-tuning on the expert-annotated dataset for each classification task, with performance assessed via cross-validation. DeepSeek-R1 was evaluated using zero-shot and few-shot prompting, in which task instructions and representative examples were included directly in the input. Prompts refined through error analysis to improve coverage of clinically similar or ambiguous subtypes. For both models, multiple independent predictions were generated and ensembled to improve robustness. All experiments were implemented using Hugging Face Transformers, a Python library for neural NLP models with standard hyperparameters.

**Figure 2.**
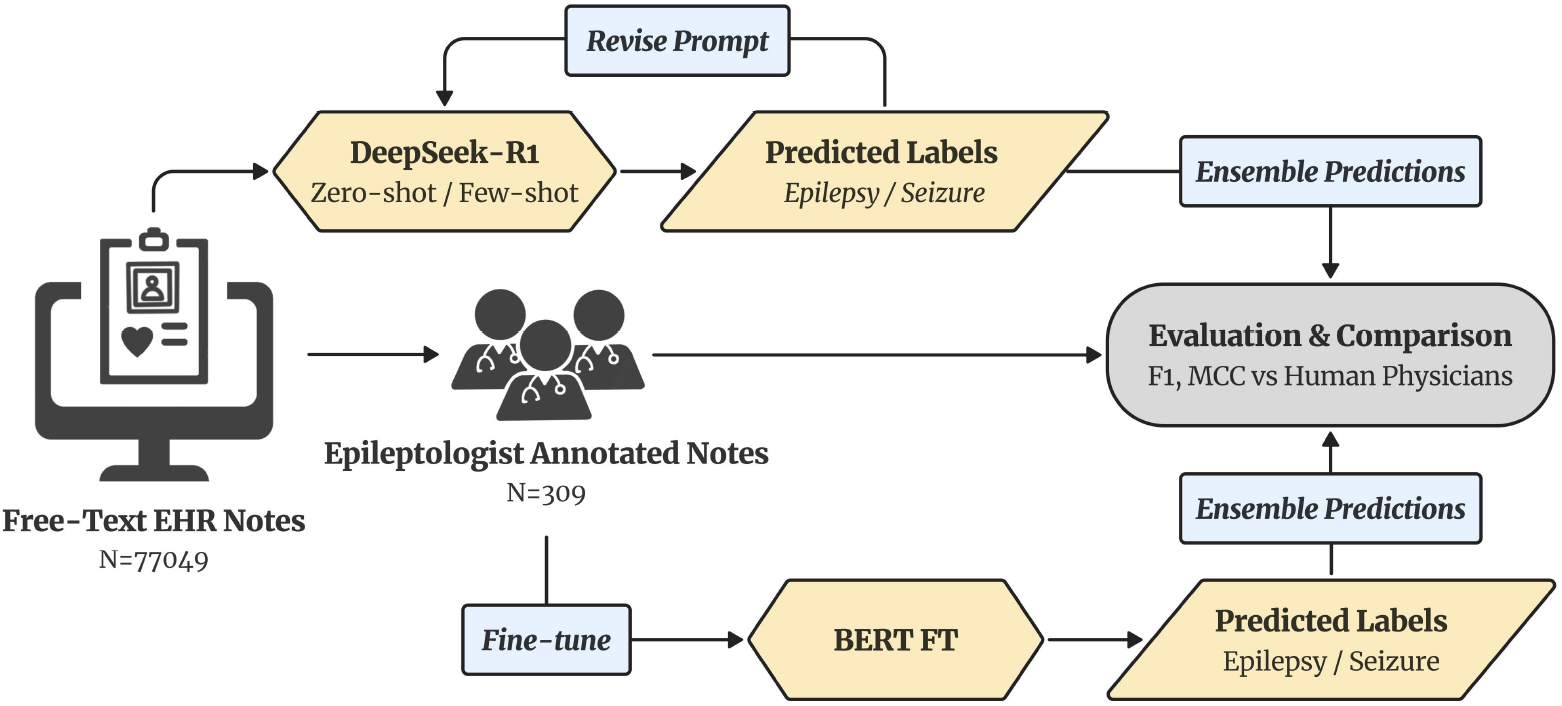
Overview of the model development pipeline. Free-text EHR notes (N=77,049) were annotated by epileptologists to create a “gold-standard” dataset (N=309). The expert-labeled dataset was used to fine-tune and evaluate the BERT model with a 70/30 training-testing split. DeepSeek-R1 was prompted in zero- and few-shot settings to predict epilepsy and seizure types directly from free-text notes. Predictions from both models were ensembled and evaluated using performance metrics to compare model performance with inter-human agreement.

After deploying the best-performing model on the full-corpus, we obtained visit-level epilepsy and seizure type labels for each progress note. Patient-level phenotypes were then derived using a two-stage adjudication: majority voting assigned a ternary epilepsy category (focal, generalized, or other) across visits, followed by selection of the most specific subtype within that category. This approach balanced longitudinal documentation variability with clinically meaningful patient-level classification.

### 2.4 Statistical Analysis

Inter-rater agreement for the single-label epilepsy-type task was assessed using Cohen’s Kappa (κ) and F1 score. For multi-label seizure type classification, agreement was assessed by treating each possible seizure type as an independent binary label, computing F1 scores for each note, then averaging these F1 scores across notes (“averaged by samples”) to obtain a single summary measure. In addition, we calculated the Jaccard similarity between the sets of seizure-type labels assigned by each annotator for each note and reported the average Jaccard score across the dataset to capture overall set-level agreement. In this multi-label setting, Jaccard similarity measures the overlap between two seizure type label sets, calculated as the size of their intersection divided by the size of their union.

Model performance was evaluated against the adjudicated ground truth using Matthews correlation coefficient (MCC) and F1 scores. MCC was used for single-label classification tasks because it provides a balanced performance measure under class imbalance, which is common in finer-grained classification. Model-to-ground-truth agreement was compared with inter-rater agreement among human annotators, with 95% confidence intervals obtained by bootstrap resampling. Binary seizure type and ternary epilepsy type classification were designated as our primary analyses, as they represent clinically meaningful distinctions with sufficient class balance.

To assess the downstream utility of automated phenotyping, we examined longitudinal diagnostic evolution and its association with seizure outcomes. We fit a continuous-time hidden Markov model (CT-HMM) using the visit-level epilepsy classifications generated by the language model to characterize longitudinal transitions between epilepsy types.^12^ The CT-HMM’s rate matrix encodes the instantaneous transition rates between diagnostic states, specifying how frequently patients move from one epilepsy classification to another per unit time. This approach accommodates irregular visit timing and differing follow-up durations. To reduce potential diagnostic drift arising from variability in documentation style, epilepsy type classifications for notes authored by minority contributors were excluded from this analysis.

After integrating the epilepsy and seizure type phenotypes with previously obtained seizure frequency and seizure freedom trajectories, we compared patient-level outcomes across epilepsy types. Seizure-free duration was summarized for each patient as a maximum seizure-free interval, and seizure burden was summarized as the average monthly seizure frequency per patient. Group differences were evaluated using the Mann-Whitney U test with effect sizes reported. Tonic-clonic seizure burden at the visit level was evaluated using a logistic generalized estimating equation with epilepsy type as the fixed effect and patient identifier as the clustering variable.

## Results

### 3.1 Interrater Reliability and Ground Truth Annotations

Label distributions of ground truth annotations are shown in **Table S1**. Inter-rater agreement (Cohen’s κ) for epilepsy types was 0.77 for ternary, 0.72 for six-way, and 0.66 for all-way classification. For seizure type annotations, κ was 0.49 for binary and 0.40 for ternary classification. For multi-label all-way seizure classification, the sample-averaged F1 score was 0.52 (SD 0.05) and the sample-averaged Jaccard similarity was 0.50 (SD 0.05), indicating that, on average, the seizure-type sets assigned by annotators overlapped by approximately 50% per note.

### 3.2 Model Performance

Performance across epilepsy and seizure type classification tasks is shown in **Figure 3**. For ternary epilepsy type classification, all models performed comparably to the benchmark of human inter-rater agreement: MCC [95% CI] for human inter-rater agreement = 0.77 [0.70-0.83], DeepSeek zero-shot ensemble = 0.85 [0.80-0.90], DeepSeek few-shot ensemble = 0.83 [0.77-0.88], and BERT ensemble = 0.73 [0.67-0.80]. Confusion matrices for the ternary epilepsy task (**Figure S1a**) show that both DeepSeek and BERT classified focal epilepsy reliably, with most errors arising from misclassification of cases in the “other” category, which is clinically more heterogenous and ambiguous. DeepSeek also maintained high accuracy for generalized epilepsy, whereas BERT demonstrated greater misclassification between generalized epilepsy and the other classes.

**Figure 3.**
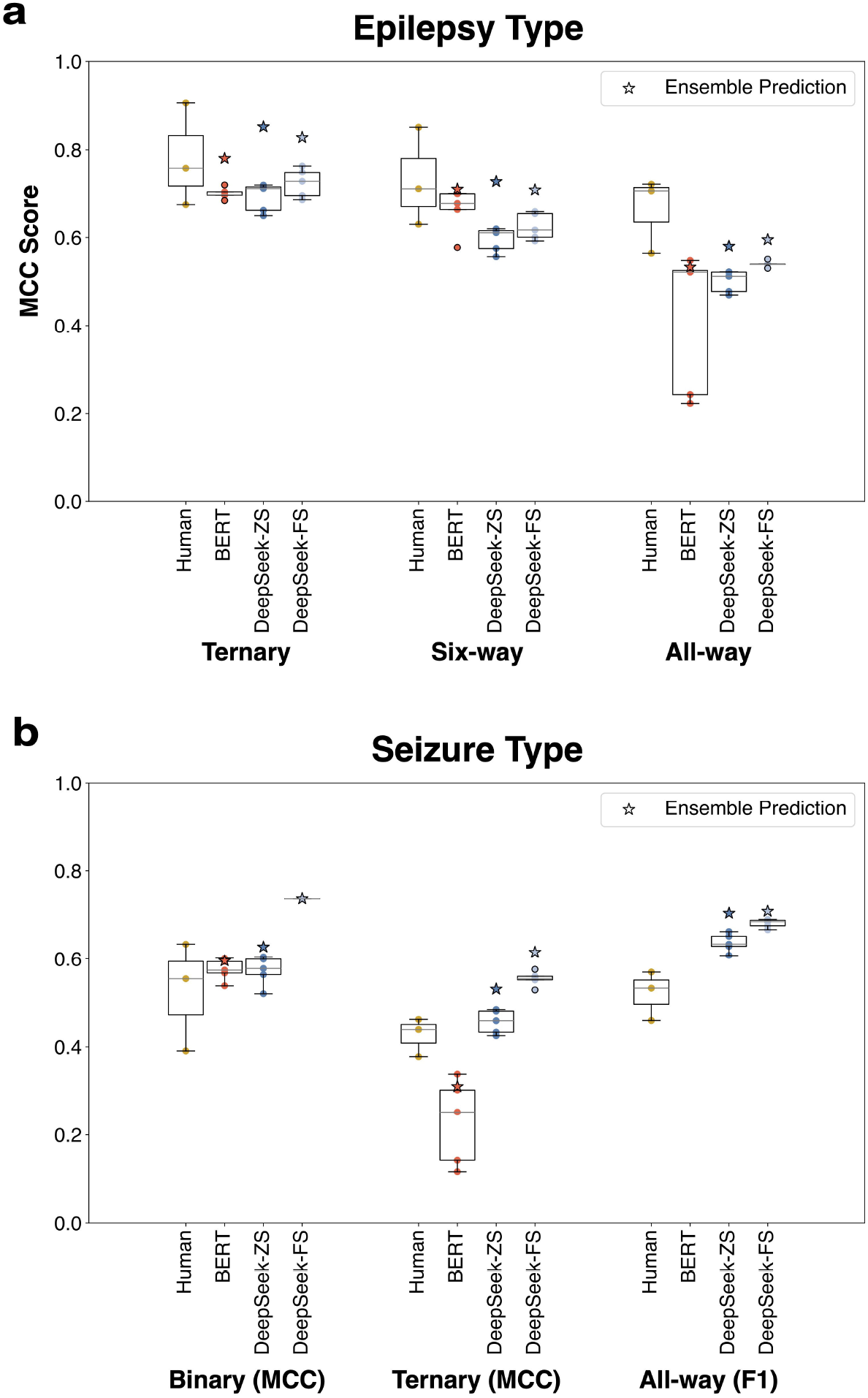
Performance comparison across epilepsy and seizure classification tasks. Boxplots represent the distribution of model performance across runs. Stars denote ensemble predictions. (a) Model performance for epilepsy type classification, evaluated under ternary, six-way, and all-way label settings. MCC scores are shown for human annotation agreement, BERT, DeepSeek-ZS, and DeepSeek-FS models. (b) Model performance for seizure type classification, evaluated under binary, ternary, and all-way label settings. Performance is reported using MCC for the binary and ternary tasks, and F1 score for multi-label all-way classification. ***Abbreviations:*** *MCC, Matthews correlation coefficient; ZS, zero-shot; FS, few-shot*

For six-way epilepsy classification, DeepSeek (zero-shot ensemble = 0.73 [0.67-0.79]; few-shot ensemble = 0.71 [0.65-0.77]) outperformed BERT (0.60 [0.53, 0.67]). Inter-annotator agreement for the six-way task (0.72, [0.65-0.78]) overlapped with both DeepSeek confidence intervals indicating model performance was comparable to human agreement.

For all-way epilepsy classification, both DeepSeek ensembles (0.58-0.60 [0.52-0.65]) and BERT (0.53 [0.43-0.64]) were lower than human inter-rater agreement (0.66 [0.60-0.71]) although the confidence intervals overlapped.

For seizure type classification, model performance consistently exceeded inter-annotator agreement across binary, ternary, and multi-label all-way tasks. For binary classification, human annotator MCC = 0.49 [0.39-0.59], DeepSeek zero-shot ensemble = 0.63 [0.54-0.71], DeepSeek few-shot ensemble = 0.74 [0.66-0.81], and BERT ensemble = 0.60 [0.49-0.69]. Confusion matrices (**Figure S1b**) show that both models correctly identified most tonic-clonic seizures, but BERT was more likely than DeepSeek to misclassify non-convulsive seizures as convulsive.

For ternary seizure classification, DeepSeek (zero-shot ensemble = 0.53 [0.45-0.61]; few-shot ensemble = 0.62 [0.54-0.69]) again outperformed human annotators (0.40 [0.31-0.48]), whereas BERT demonstrated markedly lower performance (0.31 [0.22-0.40]).

In all-way multi-label seizure classification, DeepSeek outperformed human annotators: human F1 = 0.52 [0.47-0.57], DeepSeek ensemble zero-shot F1 = 0.70 [0.67-0.74], and DeepSeek ensemble few-shot F1 = 0.72 [0.69-0.79]. In contrast, BERT failed to produce meaningful outputs for the multi-label all-way seizure-type task, instead defaulting to the same label set for all notes.

### 3.3 Full Corpus Results

We next tested the downstream applications of automated epilepsy phenotyping by applying the best-performing model to the full corpus of 77,049 clinical notes from 18,566 unique patients. We selected DeepSeek-R1 given its expert-level performance in benchmark evaluations. For epilepsy type classification, we used few-shot prompting, which provided slight improvement over zero-shot prompting with minimal additional computational cost. For seizure type classification, zero-shot prompting was used because it achieved expert-level performance with lower inference time. For both, although ensemble configurations yielded modest performance gains, we deployed a single-model configuration to enable efficient large-scale inference.

Epilepsy types for the full corpus are shown in **Table S2**. A continuous-time hidden Markov model (**Figure 4**) revealed that unspecified focal epilepsy and unclassified states have the highest outgoing transition intensities. In contrast, specified focal epilepsy and specified generalized epilepsy exhibit the highest self-transition probabilities.

**Figure 4.**
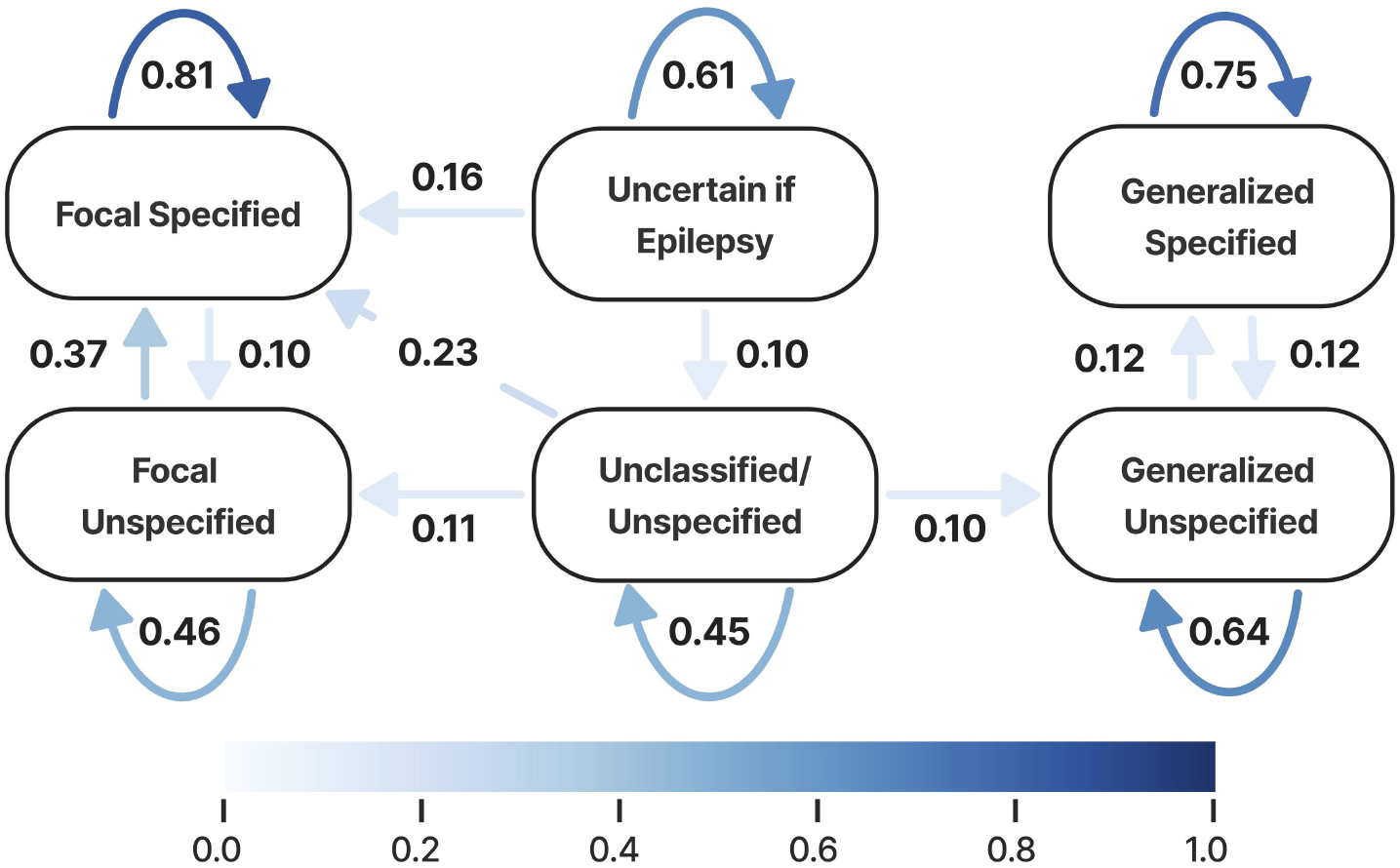
Continuous-time hidden Markov model (CT-HMM) of epilepsy type transitions over a 12-month interval. Nodes represent epilepsy classifications, and directed arrows indicate estimated transition probabilities between states. Arrow color intensity corresponds to transition probability, with darker blue indicating higher probability. Curved arrows denote self-transitions, reflecting state persistence over time. Only transitions with probability greater than 0.10 are shown.

Seizure types for the full corpus are shown in **Table S3**. Seizure type predictions demonstrated substantial overlap across categories (**Figure S2**). Tonic-clonic, focal impaired-awareness, and focal aware seizures frequently co-occurred within the same patient’s record. Many patients experienced both tonic-clonic and non-tonic-clonic events, as well as overlapping loss-of-awareness and non-loss-of-awareness seizures; psychogenic nonepileptic seizures (PNES) also often occurred in patients who have epileptic seizures rather than in isolation.

We next combined the model-derived epilepsy and seizure phenotypes with previously obtained visit-level seizure-control measures: seizure freedom and seizure frequency.^13^ Seizure freedom metrics were similar between focal and generalized epilepsies. The mean proportion of seizure-positive visits was 0.64 for focal epilepsy and 0.62 for generalized epilepsy, with no evidence of meaningful difference between groups (Mann– Whitney U test, p = 0.06; rank-biserial correlation = 0.03), consistent with the similar distribution of patients across quintiles of seizure-free visit proportion (**Figure 5a**). The maximum seizure-free duration per patient in months was also similar between epilepsy types (mean (SD) for focal = 24 (SD 23), generalized = 26 (SD 25), Mann-Whitney U test, p = 0.11; rank-biserial correlation = 0.04)

**Figure 5.**
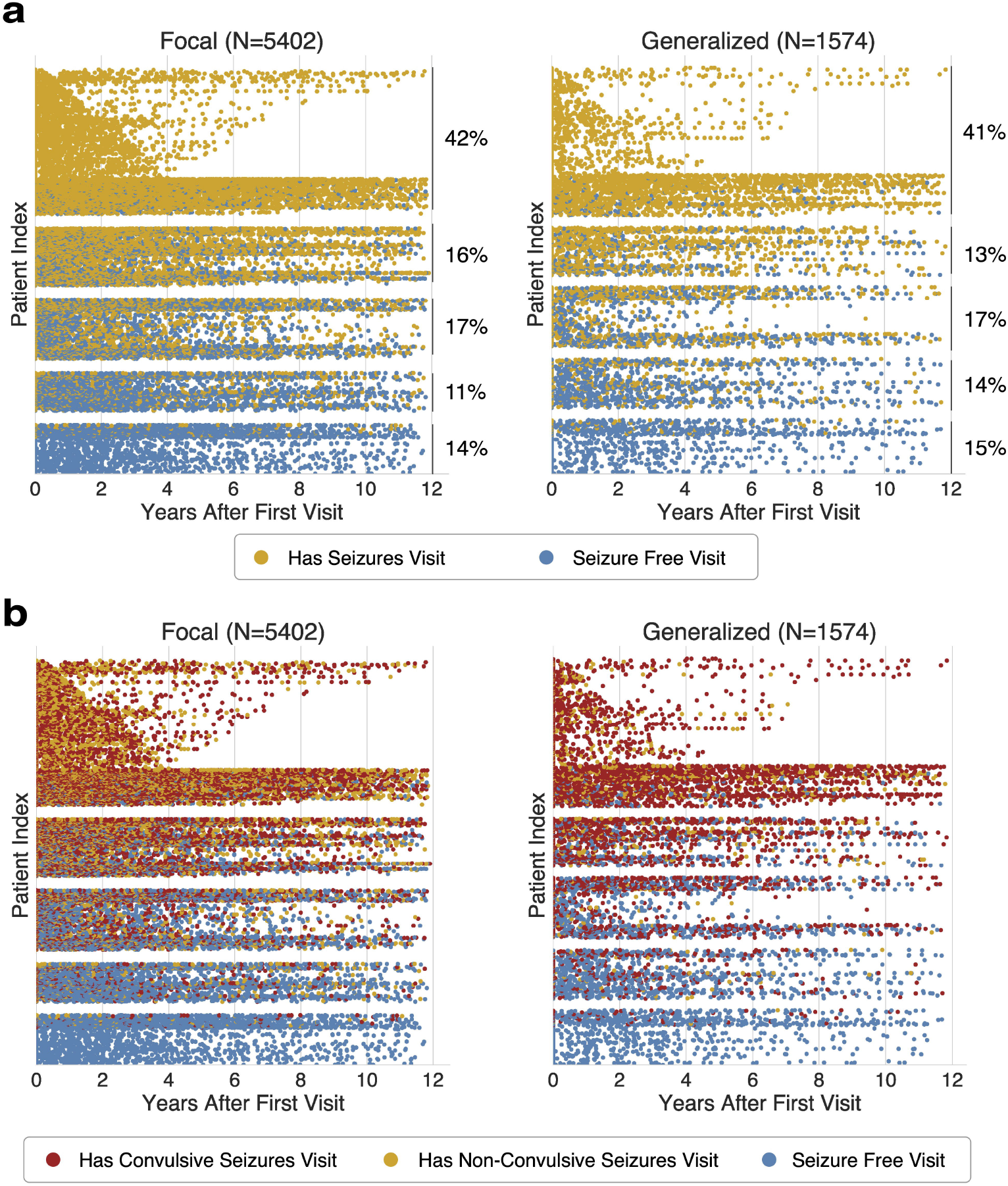
Visit-level seizure freedom trajectories stratified by model-derived phenotypes. Each dot represents a clinic visit, positioned by years after first visit (x-axis) and patient index (y-axis). Patients within each panel are grouped into horizontal bands corresponding to quintiles of the proportion of seizure-free visits (0-0.20, …, 0.80-1.00 from top down). (a) Seizure freedom aligned to each patient’s first epilepsy visit, grouped by epilepsy type: focal epilepsies (left, N=5402) and generalized epilepsies (right, N=1574). Blue dots indicate seizure-free visits and yellow dots indicate visits that are not seizure free. The proportion of patients in each quintile is displayed to the right. (b) Seizure outcomes aligned to first visit and stratified by epilepsy type. Colors denote seizure status at each visit: seizure-free (blue), non-tonic-clonic seizures (yellow), and tonic-clonic seizures (red).

In contrast, the presence of tonic-clonic seizures differed between epilepsy types (**Figure 5b**). Restricting the analysis to seizure-positive visits, tonic-clonic seizures accounted for 57% of seizure-positive visits for patients with focal epilepsy, whereas for generalized epilepsy, tonic-clonic seizures comprised 83% of seizure-positive visits. Generalized epilepsy was associated with approximately four-fold higher odds of tonic-clonic seizures compared with focal epilepsy (OR = 3.97, 95% CI [3.52-4.48], p < 0.001). The maximum seizure-free duration per patient in months was shorter for patients with tonic-clonic seizures than for those without tonic-clonic seizures (tonic-clonic patients = 23 (SD 24), non-tonic-clonic = 27 (SD 25), Mann-Whitney U test, p < 0.01; rank-biserial correlation = 0.11).

Patient-level average seizure frequency per month also differed significantly between focal and generalized epilepsy (focal = 7.0 (SD 8.4), generalized = 8.6 (SD 9.4), Mann-Whitney U test, p = 0.003) (**Figure S3**). However, the effect size was small (rank-biserial correlation = 0.075), indicating substantial overlap between the two groups. Between finer-grained epilepsy subtypes, a comparison between patients with Lennox-Gastaut syndrome (LGS) compared to all other epilepsy subtypes pooled revealed a markedly larger difference in seizure frequency, with LGS patients experiencing substantially higher seizure burdens (Mann-Whitney U test, p = 1.0×10^-17^; rank-biserial correlation = 0.40), consistent with the known severity and treatment resistance of LGS.^14^

## 4 Discussion

In this study, we show that pre-trained language models can accurately phenotype epilepsy and seizure types from unstructured clinical notes at scale, with performance comparable to – and in some tasks exceeding – expert agreement. Across classification tasks, the autoregressive model, DeepSeek-R1, consistently outperformed a fine-tuned BERT model, highlighting the advantages of LLMs for interpreting heterogeneous clinical documentation.

The deployment of the best-performing model to more than 77,000 clinical notes from over 18,500 patients yielded the largest automated phenotyping of epilepsy and seizure types to date. To contextualize these findings, we report summary patterns alongside estimates from prior large cohorts at academic epilepsy centers. In our cohort, 9,958 patients (54%) were predicted to have at least one focal seizure, similar to prior reports that focal-onset seizures account for the majority of epilepsy cases.^15^ 63% were classified as having focal epilepsy, and more than one-third of these patients had temporal lobe epilepsy. Existing studies have reported that focal epilepsies comprise roughly 60% of diagnoses, with temporal epilepsy as the most frequent subtype.^16,17^

Using these longitudinal phenotypes, our continuous-time Markov model revealed that unspecified focal epilepsy and unclassified states had the highest outgoing transition probabilities, consistent with clinical scenarios in which many patients initially receive broad or uncertain diagnoses that become more specific as EEG, imaging, and longitudinal data accumulate. In contrast, specified focal and generalized epilepsies had the highest self-transition probabilities, indicating relative diagnostic stability once a specific syndrome is established. This pattern aligns with the International League Against Epilepsy (ILAE) framework, which explicitly treats epilepsy classification as an iterative process over time.^18^ Similar diagnostic evolution over time has been described in prior cohort studies, which show that a substantial fraction of patients undergo changes in epilepsy classification as new data emerge.^19^

The substantial overlap among seizure types underscores the dynamic and heterogeneous nature of real-world epilepsy phenotypes. Many patients exhibited more than one seizure type over time, including common co-occurrence of psychogenic non-epileptic seizures and true epileptic events, reflecting both true clinical heterogeneity and the evolving characterization of events across visits.^19,20^ These findings highlight the importance of note-level phenotyping rather than relying on static, single-class labels.

Finally, integrating these phenotypes with seizure outcome measures provided insight into disease trajectories within academic epilepsy centers. Although generalized epilepsies are often considered to have a more favorable prognosis, prior work has shown that this difference is modest.^21,22^ In our cohort, maximum seizure-free durations were similar between focal and generalized epilepsies, while generalized epilepsy was associated with higher tonic-clonic seizure burden and modestly higher seizure frequency. These findings arise from an observational cohort drawn from a tertiary referral center and should not be interpreted as population-level epidemiology. Rather, they provide important insights about the phenotypes of patients at academic epilepsy centers, who are candidates for advanced clinical care and often form the basis of research cohorts.

Several limitations warrant consideration. DeepSeek inference time was substantially longer than that of fine-tuned BERT, potentially limiting real-time deployment (**Table S5**). Unlike BERT, DeepSeek-R1 prompts were evaluated on the full labeled dataset. Although no model weights were updated, iterative prompt refinement could inflate performance estimates.^23^ Future work should assess prompt robustness on fully held-out datasets from independent institutions. Next, our model was applied within a single level IV epilepsy center. Generalizability to other institutions, patient populations, documentation styles, and EHR systems will require further evaluation. Finally, although our models were benchmarked against expert-annotated data, annotator disagreement highlights that the ground truth itself contains some uncertainty.

Overall, transformer-based models extracted clinically meaningful epilepsy and seizure phenotypes at scale with accuracy comparable to epileptologists. Integrating longitudinal seizure-control metrics with model-derived phenotypes, we characterized epilepsy types and seizure outcomes across a large longitudinal cohort. This framework supports applications such as improved SUDEP risk stratification, earlier identification of candidates for surgical or device-based evaluation, and automated stratification of treatment-response cohorts. Automated phenotyping may also reduce manual abstraction by pre-populating clinical registries and trial screening lists, increasing the feasibility of large-scale, multi-site epilepsy research. Together, this work establishes a scalable approach to epilepsy phenotyping and provides reusable infrastructure for future clinical and translational studies.

## Data Availability

The data in the present study are derived from IRB-approved electronic health records containing protected health information and are not publicly available.

## Acknowledgements

EC was supported by the National Institute of Neurological Disorders and Stroke of the National Institute of Health (NIH NINDS) Award Number 2T32NS091006-11A1. KX was supported by NIH NINDS Award Number T32NS091006. ECC was supported by NIH NINDS Award Number K23 NS121401-01A1 and the Burroughs Welcome Fund. BL was supported by NIH DP1-NS-122038, and R01-NS-125137. CAE was supported by NIH NINDS Award Number K23NS121520.

