## Supplemental Appendix for "Automated epilepsy and seizure type phenotyping with pre-trained language models"

### Table of Contents


**Tables** 2

Table S1. Label distributions of ground truth annotations. 2

Table S2. Distribution of epilepsy types across full corpus. 3

Table S3. Distribution of seizure types across full corpus. 4

Table S4. Model and epileptologist performances on all epilepsy and seizure classification tasks. 5

Table S5. Model runtimes. 6

**Figures** 7

Figure S1. Confusion matrices for primary phenotyping tasks. 7

Figure S2. Patient overlap between model-derived seizure classifications. 8

Figure S3. Mean seizure frequency distributions by epilepsy types. 9

**Methods** 10

Model selection, fine-tuning, and implementation 10

Full DeepSeek-R1 Prompts 10

**References** 13

### Tables

**Table S1. Label distributions of ground truth annotations.**

For seizure type labels, because each patient could have more than one type of seizure, the total percentage across all seizure categories sum to more than 100%.

| Epilepsy & Seizure Labels | Counts |
|---|---|
| **Epilepsy Types** | |
| Focal Epilepsy | 176 (47%) |
| Generalized Epilepsy | 60 (19%) |
| Unclassified/Unspecified | 73 (24%) |
| **Seizure Types** | |
| Convulsive (Generalized Tonic-Clonic) | 217 (70%) |
| Non-Convulsive with Loss of Awareness | 157 (51%) |
| Non-Convulsive without Loss of Awareness | 224 (72%) |

**Table S2. Distribution of epilepsy types across full corpus.**
Classifications from full corpus after post-processing. We excluded notes with invalid predictions, as well as notes authored by epileptologists for patients without a suspected epilepsy diagnosis.

| Epilepsy Types Across Patients | Counts |
|---|---|
| **Focal Epilepsy** | **8498 (63%)** |
| Temporal Lobe | 3303 (24%) |
| Frontal Lobe | 1321 (10%) |
| Parietal/Occipital Lobe | 625 (5%) |
| Other Specified Focal | 1564 (11%) |
| Multifocal | 161 (1%) |
| Unlocalized Focal | 1524 (11%) |
| **Generalized Epilepsy** | **2011 (15%)** |
| Juvenile Myoclonic | 686 (5%) |
| Absence, Childhood or Juvenile | 80 (1%) |
| Other Specified Generalized | 255 (2%) |
| Unspecified Generalized | 990 (7%) |
| **Other** | **2972 (22%)** |
| Combined Generalized & Focal | 78 (1%) |
| Unclassified or Unspecified | 1700 (12%) |
| Uncertain if Epilepsy | 1055 (8%) |
| Non-Epileptic Seizure Disorder | 139 (1%) |

**Table S3. Distribution of seizure types across full corpus.**
Percentages do not sum to 100% because individual patients can be assigned more than one seizure type. We excluded notes with invalid predictions, as well as notes authored by epileptologists for patients without a suspected epilepsy diagnosis.

| Seizure Types Across Patients | Counts |
|---|---|
| **Focal Aware** | 6484 (35%) |
| **Focal Impaired Awareness** | 7302 (39%) |
| **Focal Unspecified** | 1831 (10%) |
| **Absence** | 1171 (6%) |
| **Convulsive, Tonic-Clonic** | 9571 (52%) |
| **Drop Attack, Atonic** | 558 (3%) |
| **Myoclonic** | 1842 (10%) |
| **Tonic** | 243 (1%) |
| **Clonic** | 264 (1%) |
| **Unspecified Staring Spell** | 1061 (6%) |
| **Unspecified** | 6170 (33%) |
| **Psychogenic Non-Epileptic** | 1787 (10%) |

**Table S4. Model and epileptologist performances on all epilepsy and seizure classification tasks.** The highest metrics per task are highlighted in bold.

| Task | Model | Weighted-F1 | Macro-F1 | MCC |
|---|---|---|---|---|
| **Ternary Epilepsy Type** (Focal/Generalized/ Other) | Epileptologists | 0.86 | 0.86 | 0.77 |
| | BERT | 0.82 (SD 0.008) | 0.78 (SD 0.011) | 0.69 (SD 0.01) |
| | DeepSeek ZS | 0.84 (SD 0.02) | 0.61 (SD 0.01) | 0.69 (SD 0.03) |
| | DeepSeek FS | 0.86 (SD 0.02) | 0.64 (SD 0.02) | 0.731 (SD 0.03) |
| | BERT-Ensemble | 0.85 | 0.81 | 0.73 |
| | **DeepSeek ZS-Ensemble** | **0.91** | **0.89** | **0.85** |
| | DeepSeek FS-Ensemble | 0.89 | 0.87 | 0.83 |
| **Six-way Epilepsy Type** (Focal/Gen/ Combined/ Unclassified, Unspecified/ Epilepsy Uncertain/ PNES) | Epileptologists | 0.82 | 0.72 | 0.72 |
| | BERT | 0.68 (SD 0.06) | 0.38 (SD 0.04) | 0.52 (SD 0.09) |
| | DeepSeek ZS | 0.75 (SD 0.01) | 0.50 (SD 0.007) | 0.59 (SD 0.03) |
| | DeepSeek FS | 0.78 (SD 0.02) | 0.53 (SD 0.03) | 0.63 (SD 0.03) |
| | BERT-Ensemble | 0.73 | 0.40 | 0.60 |
| | **DeepSeek ZS-Ensemble** | **0.81** | 0.68 | **0.73** |
| | **DeepSeek FS-Ensemble** | 0.81 | **0.71** | 0.71 |
| **All-way Epilepsy Type** | Epileptologists | 0.71 | 0.61 | 0.66 |
| | BERT | 0.42 (SD 0.2) | 0.29 (SD 0.1) | 0.41 (SD 0.1) |
| | DeepSeek ZS | 0.58 (SD 0.02) | 0.52 (SD 0.01) | 0.50 (SD 0.02) |
| | DeepSeek FS | 0.63 (SD 0.005) | 0.54 (SD 0.01) | 0.54 (SD 0.007) |
| | BERT-Ensemble | 0.56 | 0.37 | 0.53 |
| | DeepSeek ZS-Ensemble | 0.64 | 0.56 | 0.58 |
| | **DeepSeek FS-Ensemble** | **0.66** | **0.58** | **0.60** |
| **Binary Seizure Type** (Convulsive/Non-Convulsive) | Epileptologists | 0.74 | 0.74 | 0.49 |
| | BERT | 0.82 (SD 0.05) | 0.78 (SD 0.06) | 0.58 (SD 0.1) |
| | DeepSeek ZS | 0.80 (SD 0.02) | 0.78 (SD 0.02) | 0.57 (SD 0.03) |
| | DeepSeek FS | 0.85 (SD 0.002) | 0.83 (SD 0.002) | 0.67 (SD 0.005) |
| | BERT-Ensemble | 0.83 | 0.79 | 0.60 |
| | DeepSeek ZS-Ensemble | 0.83 | 0.80 | 0.63 |
| | **DeepSeek FS-Ensemble** | **0.88** | **0.86** | **0.74** |
| **Ternary Seizure Type** (Convulsive, LOA/Non-Convulsive, LOA/ Non-Convulsive, No LOA) | Epileptologists | 0.64 (SD 0.01) | 0.58 (SD 0.04) | 0.43 (SD 0.04) |
| | BERT | 0.61 (SD 0.06) | 0.44 (SD 0.04) | 0.23 (SD 0.09) |
| | DeepSeek ZS | 0.73 (SD 0.01) | 0.58 (SD 0.02) | 0.46 (SD 0.02) |
| | DeepSeek FS | 0.78 (SD 0.009) | 0.65 (SD 0.02) | 0.55 (SD 0.02) |
| | BERT-Ensemble | 0.68 | 0.48 | 0.31 |
| | DeepSeek ZS-Ensemble | 0.77 | 0.63 | 0.53 |
| | **DeepSeek FS-Ensemble** | **0.82** | **0.69** | **0.62** |
| | | **Samples-F1** | **Macro-F1** | **Exact Match** |
| **All-way Seizure Type** | **Epileptologists** | 0.52 | 0.55 | **0.46** |
| | BERT | NA | NA | NA |
| | **DeepSeek ZS** | 0.64 (SD 0.02) | 0.52 (SD 0.04) | 0.33 (SD 0.006) |
| | DeepSeek FS | 0.68 (SD 0.009) | 0.56 (SD 0.04) | 0.40 (SD 0.001) |
| | DeepSeek ZS-Ensemble | 0.70 | 0.56 | 0.37 |
| | **DeepSeek FS-Ensemble** | **0.72** | **0.58** | 0.37 |

**Table S5. Model runtimes.**
Mean (± standard deviation when available) on epilepsy and seizure classification tasks.

| | | **Epilepsy Type** | | **Seizure Type** | |
|---|---|---|---|---|---|
| **Phase** | **Metric** | BERT | DeepSeek | BERT | DeepSeek |
| **Training** | **Throughput** (samples/sec) | 17.4 (±1.3) | NA | 16.7 (± 0.1) | NA |
| **Testing** | **Throughput** (samples/sec) | 53.1 (± 5.2) | 0.05 (± 0.006) | 51.3 (± 0.4) | 0.04 (± 0.004) |
| **Testing** | **Latency** (sec/sample) | 0.02 (±0.002) | 21.7 (± 3.3) | 0.02 (± 0.0002) | 26.7 (± 2.7) |
| **Full-Corpus** | **Latency** (sec/sample) | NA | 14.0 | NA | 24.7 |

### Figures

**Figure S1. Confusion matrices for primary phenotyping tasks.**
(A) Ternary epilepsy type classification and (B) binary seizure type classification tasks.

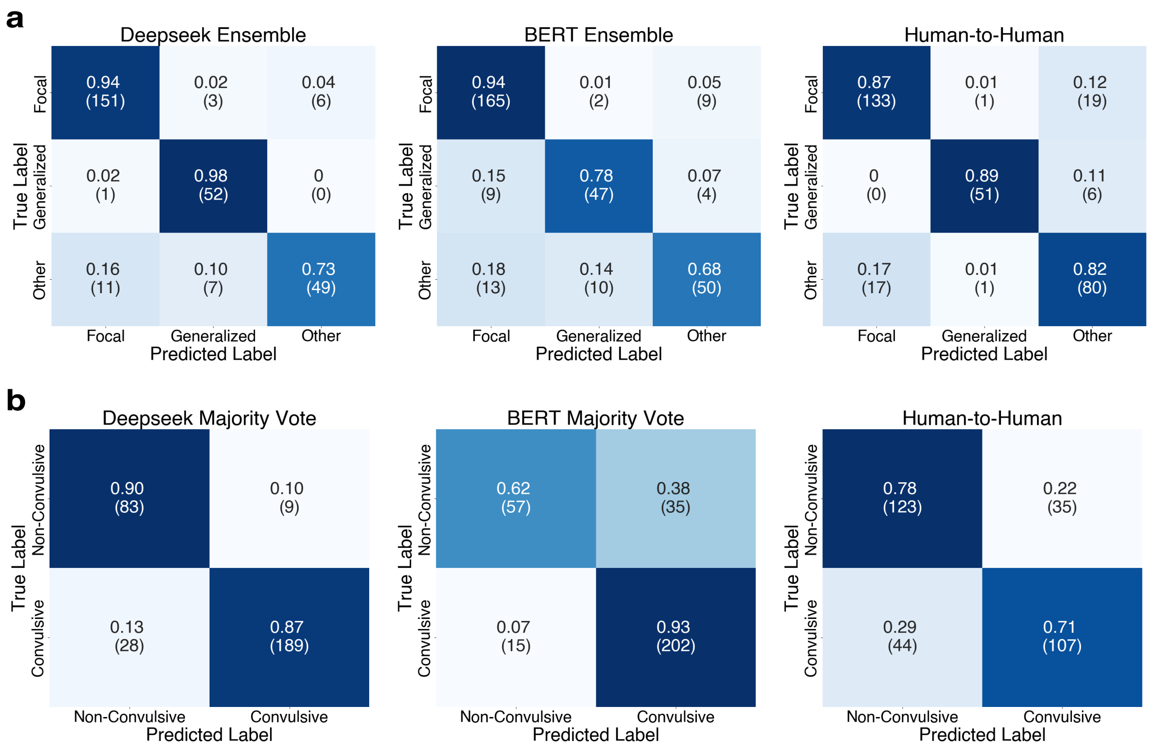

**Figure S2. Patient overlap between model-derived seizure classifications.**
Venn diagrams show the overlap in seizure type assignments at the patient level. Counts indicate the number of patients shared between seizure categories. Abbreviations: FIAS, focal impaired awareness seizure; FAS, focal aware seizure; PNES, Psychogenic Non-Epileptic Seizures; LOA, Loss of Awareness.

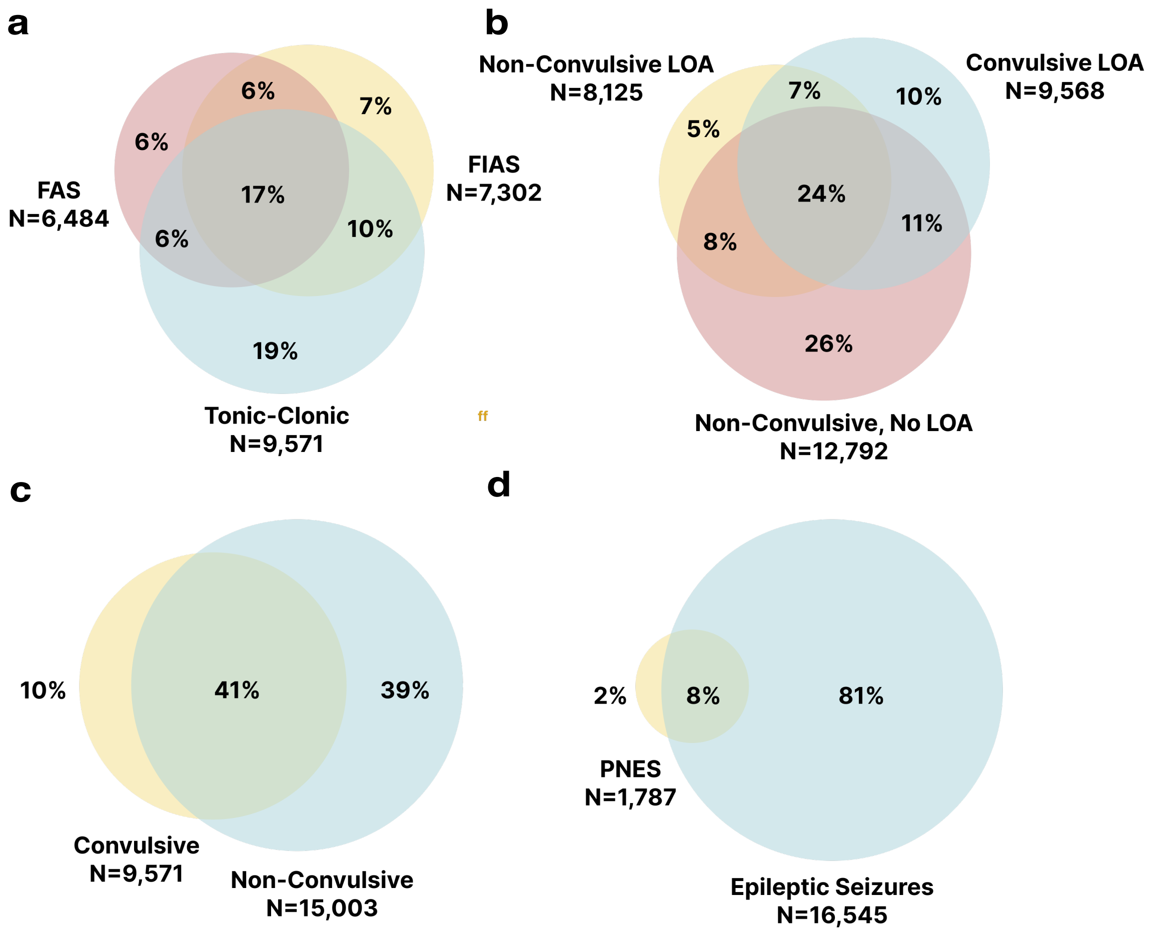

**Figure S3. Mean seizure frequency distributions by epilepsy types.**
Distribution of patient-level seizure frequency (seizures per month) by epilepsy subtype. Violin plots depict the full distribution of seizure frequency values within each epilepsy type, with embedded box plots indicating the median and interquartile ranges.

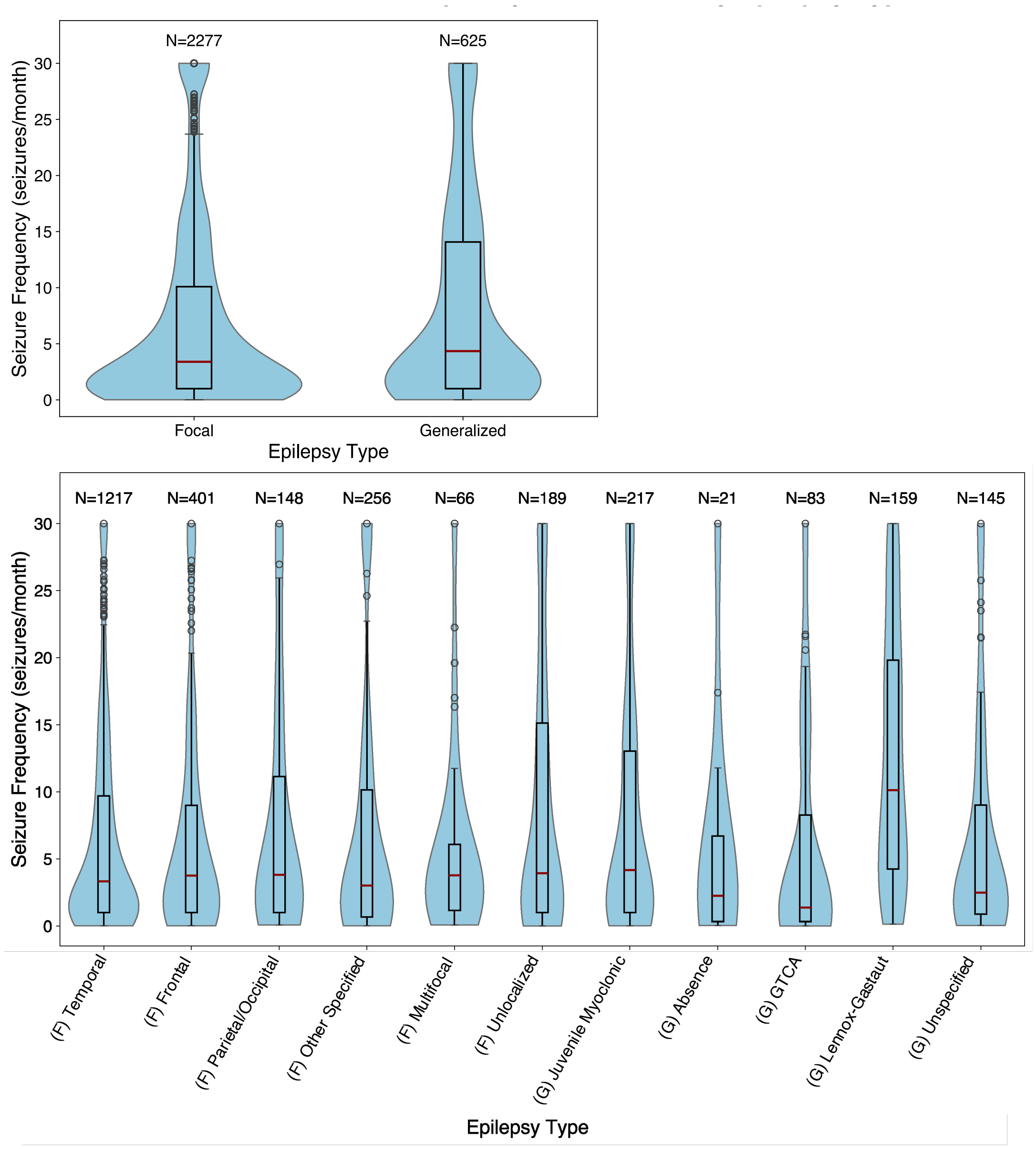

### Methods

**Model selection, fine-tuning, and implementation**

BERT (specifically the newer BGE variant[1]) was selected over BERT's bioclinical counterparts (e.g., Bio_ClinicalBERT) because prior evaluations on similar epilepsy-related information extraction tasks showed no meaningful performance gains from domain-specific variants.[2] DeepSeek-R1 was chosen over other autoregressive models because its training driven by reinforcement learning has been shown to elicit stronger emergent reasoning capabilities, particularly for problem solving in STEM-related fields, while also offering publicly available model weights that make it suitable for clinical research applications.[3,4]

For BERT, we first performed unsupervised domain adaptation with masked language modeling using ~64,000 unannotated progress notes. We then performed task-specific fine-tuning using publicly available English datasets, BoolQ and BoolQ3L for the binary and ternary classification tasks.[5,6] Finally, we performed supervised fine-tuning with training data from the adjudicated "ground truth" datasets, tailoring the model to each classification task. To evaluate BERT on our dataset, we used 3-fold cross-validation, ensuring that model performance was assessed across multiple splits of the annotated data for more robust estimates.

For DeepSeek-R1, we used a systematic prompting workflow under both zero-shot and few-shot configurations. In the zero-shot setting, prompts contained only task instructions without examples, while in the few-shot setting, we supplemented instructions with a small number of representative input-output pairs. These prompts were iteratively refined through cycles of testing and analysis: we applied candidate prompts to a sample of labeled clinical notes, performed error analysis to identify recurring issues (e.g., confusion between clinically similar subtypes), and revised the prompts to improve clarity and coverage (e.g., modifying examples, refining task wording). This process was repeated until performance on validation subsets stabilized, suggesting diminishing returns from further refinements. Final prompts were applied to the full labeled dataset without further tuning. For both models, we generated five independent sets of predictions and ensembled them to improve robustness and generalizability.

All experiments were implemented using Hugging Face Transformers, a Python library for neural NLP models. For fine-tuning, we used Hugging Face's default hyperparameters and set BERT's maximum sequence length to 512 tokens to capture as much context as possible. For DeepSeek, we used the DeepSeek-R1-Distill-Llama-8B model via the Hugging Face text-generation pipeline. Inference was performed with a batch size of 32 and a maximum generation length of 4096 tokens. Each input prompt combined a fixed prefix, the note text, and a task-specific prompt with or without a few labeled examples.

**Full DeepSeek-R1 Prompts**

**A. Epilepsy Type Classification Prompt:**

"You are an epileptologist tasked with determining the type of epilepsy a patient has based on a provided clinical note. Epilepsy is a neurological disorder characterized by unprovoked and/or uncontrollable seizures. Use the following definitions to help you make your decision.

F1-F6 are all subtypes of focal epilepsy. A patient with focal epilepsy may have seizures that originate from one part of the brain and are focal seizure types (FAS/SPS, FIAS/CPS). If EEG is mentioned, it may show focal epileptiform discharges, and if brain MRI is mentioned, it may show structural abnormalities, though it may also be normal. Focal epilepsy may be associated with TBI, stroke, tumors, etc. Focal epilepsy is also referred to as 'localization-related epilepsy' in the note or other similar terminology.

F1 - Temporal lobe epilepsy: this patient's seizures were localized to a temporal lobe.

F2 - Frontal lobe epilepsy: this patient's seizures were localized to their frontal lobe.

F3 - Parietal/Occipital lobe epilepsy: this patient's seizures were localized to a parietal or occipital lobe.
F4 - Other specified focal epilepsy: this patient's seizures were explicitly localized in the note, but is not F1, F2, or F3.
F5 - Multifocal: seizures have two or more independent localizations.
F6 - Unlocalized focal epilepsy: the patient's seizures were not explicitly localized to a specific brain area in the clinical note.
G1-G5 are all subtypes of generalized epilepsy. A patient with generalized epilepsy may have generalized seizure types and generalized epileptiform discharges on EEG (generalized spike-wave) if EEG is mentioned in the note. If there is a brain MRI mentioned in the clinical note it is usually normal.
G1 - Juvenile Myoclonic Epilepsy (JME): childhood/adolescent generalized, myoclonic, and absence seizures. Note will usually explicitly specify the patient has JME.
G2 - Absence Epilepsy, Childhood or Juvenile (CAE/JAE): childhood/adolescent absence seizures.
G3 - Generalized Tonic-Clonic Seizures Alone (GTCSA): typically late-adolescent to early adult seizures that typically occur soon after waking.
G4 - Lennox-Gastaut Syndrome: the patient has Lennox-Gastaut Syndrome (LGS). The note will usually explicitly specify the patient has LGS.
G5 - Unspecified generalized epilepsy: the author of the clinical note was unsure or did not specify the exact type of generalized epilepsy.
Other types:
O1 - Combined Generalized and Focal Epilepsy: The patient has both focal and generalized epilepsies. This is not to be mistaken with the patient simply having had both focal and generalized onset seizure types.
O2 - Unspecified/Unclassified Epilepsy: The patient has unclassified/unspecified epilepsy if the note author does not provide any information or even speculate as to whether the patient may have focal or generalized epilepsy.
O3 - Uncertain if Epilepsy: It is uncertain if the patient even has epilepsy at all. Note may not mention any previous epilepsy diagnosis and patient may not have a sufficient history of seizures to diagnose them with epilepsy.
O4 - Non-Epileptic Seizure Disorder: Patient has non-epileptic seizure disorder with solely non-epileptic events (e.g., PNEEs, febrile seizures). This category means the patient does not have epilepsy.

RESPONSE FORMAT:
Format your final answer in JSON format with the following keys:
- epilepsy_type: F1, F2, F3, F4, F5, F6, G1, G2, G3, G4, G5, O1, O2, O3, O4
- reasoning: your strongest argument for why your chosen answer is the appropriate classification for the clinical note. If you encounter inconsistencies or problems in classification, work through them step by step.

IMPORTANT RULES FOR CLASSIFICATION:
Before selecting a classification, do not do the following:
1. Do not use seizure descriptions or symptoms as a basis for classifying the patient's epilepsy type.
2. Do not determine the type of epilepsy the patient has solely based on the types of seizures they have.
3. Do not make assumptions that are not made by the author in the clinical note.
4. Do not use prior knowledge about medications or what treatments the patient was given to determine the epilepsy type.
5. The clinical note may not necessarily provide definitive evidence (e.g., EEG or MRI report) for an epilepsy type, but that does not mean you cannot pick that epilepsy type. For each type, put forth your best argument

for why the patient may have that type of epilepsy. Pick the epilepsy type that you can make the strongest argument for."

**B. Seizure Type Classification Prompt:**

"You are an epileptologist who needs to identify what types of seizure events are described in the clinical note. There may be more than one seizure event type in the note. For each event described, determine which of the following seizure event types is the best fit.
F1 - Focal Aware Seizure (FAS) or Simple Partial Seizure (SPS)
F2 - Focal Impaired Awareness Seizure (FIAS) or Complex Partial Seizure (CPS)
F3 - Focal Unspecified Seizure
G1 - Absence
G2 - Clonic
G3 - Convulsions, Grand Mal, Bilateral Tonic-Clonic (BTC), Generalized Tonic-Clonic (GTC), Tonic-Clonic
G4 - Drop attack, Atonic
G5 - Myoclonic
G6 - Tonic
O1 - Unspecified Staring Spell or Petit Mal
O2 - Unspecified: the note author was unsure of or did not specify the type of seizure event the patient had
O3 - Psychogenic Non-Epileptic Seizures (PNES): Non-epileptic seizures are events similar to Epileptic Events but ultimately have no electrographic correlate. Patient may be diagnosed with PNES.
The types above may be referred to in the clinical note by their acronyms alone (e.g., CPS, GTC, PNES, etc.)

RESPONSE FORMAT
Format your final answer in JSON format with the following keys:
- seizure_types: list of seizure types picked from the following list: F1, F2, F3, G1, G2, G3, G4, G5, G6, O1, O2, O3
- reasoning: your strongest argument for why your chosen seizure types are appropriate for the clinical note. If you encounter inconsistencies or problems in classification, work through them step by step.

IMPORTANT RULES FOR CLASSIFICATION:
Before selecting a classification, do not do the following:
1. Do not make assumptions that are not made by the author in the clinical note.
2. Do not use prior knowledge about medications or what treatments the patient was given to determine the seizure type."
